# Prevalence of COVID-19-related risk factors and risk of severe influenza outcomes in cancer survivors: a matched cohort study using linked English electronic health records data

**DOI:** 10.1101/2020.10.08.20209304

**Authors:** Helena Carreira, Helen Strongman, Maria Peppa, Helen I McDonald, Isabel dos-Santos-Silva, Susannah Stanway, Liam Smeeth, Krishnan Bhaskaran

**Author notes:** equal contribution (Helena Carreira, Helen Strongman). Correspondence to: Helena Carreira, London School of Hygiene & Tropical Medicine., Keppel Street, WC1 7HT London, UK.

## Abstract

**Background:** People with active cancer are recognised as at risk of COVID-19 complications, but it is unclear whether the much larger population of cancer survivors is at elevated risk. We aimed to address this by comparing cancer survivors and cancer-free controls for (i) prevalence of comorbidities considered risk factors for COVID-19; and (ii) risk of severe influenza, as a marker of susceptibility to severe outcomes from epidemic respiratory viruses.

**Methods:** We included survivors (≥1 year) of the 20 most common cancers, and age, sex and general practice-matched cancer-free controls, derived from UK primary care data linked to cancer registrations, hospital admissions and death registrations. Comorbidity prevalences were calculated 1 and 5 years from cancer diagnosis. Risk of hospitalisation or death due to influenza was compared using Cox models adjusted for baseline demographics and comorbidities.

**Findings:** 108,215 cancer survivors and 523,541 cancer-free controls were included. Cancer survivors had more asthma, other respiratory, cardiac, diabetes, neurological, renal, and liver disease, and less obesity, compared with controls, but there was variation by cancer site. There were 205 influenza hospitalisations/deaths, with cancer survivors at higher risk than controls (adjusted HR 2.78, 95% CI 2.04-3.80). Haematological cancer survivors had large elevated risks persisting for >10 years (HR overall 15.17, 7.84-29.35; HR >10 years from cancer diagnosis 10.06, 2.47-40.93). Survivors of other cancers had evidence of raised risk up to 5 years from cancer diagnosis only (HR 2.22, 1.31-3.74).

**Interpretation:** Risks of severe COVID-19 outcomes are likely to be elevated in cancer survivors. This should be taken into account in policies targeted at clinical risk groups, and vaccination for both influenza, and, when available, COVID-19, should be encouraged in cancer survivors.

**Funding:** Wellcome Trust, Royal Society, NIHR.

**Research in context:** *Evidence before this study:* Few data are available to date on how COVID-19 affects cancer survivors. We searched PubMed with the keywords “influenza cancer survivors” to identify studies that compared severe influenza outcomes in cancer survivors and in a control group. No study was identified.

*Added value of this study:* In this matched cohort study of routinely collected electronic health records, we demonstrated raised risks of influenza hospitalisation or mortality in survivors from haematological malignancies for >10 years after diagnosis, and in survivors from solid cancers up to 5 years after diagnosis.

*Implications of all the available evidence:* Cancer survivorship appears to be an important risk factor for severe influenza outcomes, suggesting that cancer survivors may also be at raised risk of poor COVID-19 outcomes. This should be taken into account in public health policies targeted at protecting clinical risk groups. Influenza vaccination should be encouraged in this group, and may need to be extended to a wider population of medium- to long-term cancer survivors than currently recommended.

## Introduction

As of 7 July 2020, the novel Coronavirus disease 2019 (COVID-19) has been diagnosed in over 11.6 million individuals with more than 539,000 deaths reported worldwide.^1^ Around 20% of individuals contracting the virus are estimated to develop severe disease requiring hospitalisation, with a high risk of mortality.^2^ A key aspect of managing the impacts of the pandemic is understanding who is vulnerable to experiencing severe outcomes, so that mitigation strategies can be targeted at those most in need. Those under current treatment for cancer were recognised early on as being a high risk group,^3^ but the extent to which the much larger population of medium- to long-term cancer survivors, might be considered vulnerable is unknown. In England alone, this group includes over 1.8 million people.^4^

Current guidance on who should be considered vulnerable has been largely based on policies developed for previous epidemic respiratory viruses, notably influenza. For example, vaccination against influenza is only recommend for individuals under active treatment for cancer and for up two years following some treatments and haematological cancers,^5^ while longer-term cancer survivors with no recent immunosuppressing treatment are not considered high-risk in vaccination guidance from Public Health England or the American Cancer Society.^5,6^ Yet medium- to long-term cancer survivors could plausibly be at raised risk of severe COVID-19 outcomes. Studies among women with breast cancer have found chemotherapy to be associated with changes in immune parameters that did not return to pre-treatment levels a year or more after end of treatment, raising the possibility of a long-term weakened immune system in cancer survivors.^7,8^ In addition, cancer survivors have known raised risks of heart disease,^9^ which is itself an emerging risk factor for COVID-19 mortality.^10^ One large UK study identified raised risks of COVID-19 mortality in survivors of haematological malignancies even several years after cancer diagnosis,^11^ but there is little other evidence to date to inform policy around managing COVID-19 related risks in cancer survivors.

We therefore aimed to investigate whether cancer survivors are likely to be a high-risk group for severe outcomes during the current COVID-19 pandemic in two ways: first, by comparing the prevalence of risk factors currently used to guide COVID-19 policy between site-specific cancers survivors and cancer free controls; second, by comparing the risk of influenza hospitalisation or death between cancer survivors and cancer free controls, as a way of exploring susceptibility to severe outcomes from epidemic respiratory viruses.

## Methods

This study is reported as per the Strengthening the Reporting of Observational Studies in Epidemiology (STROBE) guideline (S1 Checklist).

### Study design and data sources

We carried out a population-based cohort study among 1-year survivors of the 20 most common site-specific cancers matched to cancer-free controls. We used primary care data from Clinical Practice Research Datalink (CPRD GOLD)^12^ linked to national data on hospital admissions from the Hospital Episode Statistics Admitted Patient Care (HES APC) database,^13^ cancer registrations from the National Cancer Registration and Analysis Service (NCRAS),^14^ death registrations – including cause of death information – from the Office of National Statistics mortality database, and postcode-based index of Multiple Deprivation data.^9^ CPRD GOLD comprises routinely collected clinical and administrative data from general practices in the UK that use Vision software and have chosen to participate; approximately 7% of the UK population is included. Data include Read-coded diagnoses and care events, drug prescriptions, numerical measurements (e.g., height and weight), laboratory test results (e.g. serum creatinine) and health risk factors (e.g. smoking status). Secondary care diagnoses reported to the general practitioner (GP) through discharge letters are typically recorded in the general practice record if they are considered to affect the ongoing care of the patient. Linked International Classification of Diseases, version 10 (ICD-10) coded HES APC and NCRAS data improve ascertainment of diseases treated in secondary care.^9^ Use of linked data restricted our study to England and the study period covered by all linked data sources, January 1 1990 to December 31 2015.

### Study population

Cohorts of adult cancer survivors (aged ≥18 years) were identified for each of the 20 most common cancer sites (listed in Table 1), as in a previous study.^9^ Briefly, we used CPRD GOLD, HES APC, and NCRAS to identify 1-year survivors of incident cancer diagnoses. Incident diagnoses were defined as the earliest record of a malignant cancer of interest among individuals with at least 1 year of follow-up meeting CPRD internal quality control criteria prior to the diagnosis (to ensure that the cancer was incident). The derivation of the final analysis cohort is described in Supplementary Figure S1. Cancer survivors with missing data on smoking (5.5%), body mass index (13.0%), or index of multiple deprivation (an area-based proxy for socioeconomic status derived from the patient’s postcode; <0.1%) were excluded from the cohorts.

**Table 1.** Characteristics of the patients included in analyses.

|  | **Cancer survivors** | **Comparison group** |
| --- | --- | --- |
| **Number (%)** | 108,215 (100) | 523,541 (100) |
| **Cancer site, N (%)** |  |  |
| Bladder | 7,712 (7.13) | - |
| Breast | 25,633 (23.69) | - |
| Cervix | 1,209 (1.12) | - |
| Central nervous system | 906 (0.84) | - |
| Colorectal | 14,216 (13.14) | - |
| Gastric | 1,507 (1.39) | - |
| Kidney | 2,197 (2.03) | - |
| Leukaemia | 3,419 (2.03) | - |
| Liver | 554 (0.51) | - |
| Lung | 5,369 (4.96) | - |
| Malignant melanoma | 7,098 (6.56) | - |
| Multiple myeloma | 1,843 (1.70) | - |
| Non-Hodgkin lymphoma | 4,423 (4.09) | - |
| Oesophagus | 1,794 (1.66) | - |
| Oral cavity | 1,584 (1.46) | - |
| Ovary | 2,710 (2.50) | - |
| Pancreas | 864 (0.80) | - |
| Prostate | 20,709 (19.14) | - |
| Thyroid | 1,028 (0.95) | - |
| Uterus | 3,440 (3.18) | - |
| **Years from index date to end of follow-up** |  |  |
| Mean (SD) | 5.7 (4.7) | 7.0 (4.7) |
| Median (IQR) | 4.7 (1.9, 8.4) | 6.2 (3.3, 9.9) |
| Range | 0.0-24.2 | 0.0-24.2 |
| **Total person-years included (thousands)** | 620.144 | 3666.849 |
| **Demographics** |  |  |
| **Age at index date (years)** |  |  |
| Mean (SD) | 66.1 (13.3) | 66.0 (13.2) |
| Median (IQR) | 67.0 (58.0, 76.0) | 67.0 (58.0, 76.0) |
| **Age at index date (years)** |  |  |
| 18-39 | 4,028 (3.7) | 19,208 (3.7) |
| 40-59 | 27,029 (25.0) | 131,049 (25.0) |
| 60-79 | 60,347 (55.8) | 293,080 (56.0) |
| >=80 | 16,811 (15.5) | 80,204 (15.3) |
| **Gender** |  |  |
| Men | 51,541 (47.6) | 245,760 (46.9) |
| Women | 56,674 (52.4) | 277,781 (53.1) |
| **IMD quintile** |  |  |
| 1 (least deprived) | 19,334 (17.9) | 94,233 (18.0) |
| 2 | 21,439 (19.8) | 103,694 (19.8) |
| 3 | 20,649 (19.1) | 99,684 (19.0) |
| 4 | 23,114 (21.4) | 111,768 (21.3) |
| 5 (most deprived) | 23,679 (21.9) | 114,162 (21.8) |
| **Previous vaccination at index date** |  |  |
| Influenza | 72,924 (67.4) | 322,908 (61.7) |
| Pneumococcal | 48,953 (45.2) | 224,496 (42.9) |

Older records were more likely to be excluded, as completeness of lifestyle information improved when the Quality and Outcomes Framework was introduced in 2004.^12^ Cancer survivors were followed up from 1 year after diagnosis (index date) and matched on age (±3 years), sex, and general practice to 5 controls with no history of cancer and at least 2 years of follow-up prior to the index date of the matched cancer survivor (since cancer survivors had to have one year of follow-up before and after the date of cancer diagnosis to be included). Cancer survivors were eligible to be selected as controls until the date of the incident cancer.

### Outcome and covariates

The main outcome for the study was influenza hospitalisation or death, identified using ICD-10 codes in HES and ICD-9 and ICD-10 codes in ONS mortality data (codes available in Supplementary Table S1). For the primary analysis, we counted hospitalisations with a primary diagnosis of influenza, and deaths with an underlying cause of influenza. In a sensitivity analysis, we broadened the definition to include hospitalisations/deaths with any code for influenza present.

Age and sex were matching factors. Other covariates were index of multiple deprivation quintile, smoking status (never, former, current smoker), and common comorbidities identified *a priori* as of potential importance in determining risk of severe COVID-19 outcomes, namely asthma, chronic respiratory disease other than asthma, chronic heart disease, chronic liver disease, chronic neurological disease, chronic kidney disease, diabetes, obesity, sickle cell disease or splenectomy. Other causes of immunosuppression were not included due to overlap with cancer and its treatment. In a secondary analysis we also described the total number of comorbidities (0 vs 1 vs ≥2 comorbidities from the aforementioned list). Full variable definitions and code lists are provided in Supplementary Table S1.

### Statistical analysis

#### Prevalence of COVID-19 related risk factors in cancer survivors and controls

Among cancer survivors and controls alive and under follow-up in CPRD GOLD at the index date (i.e. 1 year after cancer diagnosis for cancer survivors) and 4 years later (5 years after diagnosis), we calculated the proportion with each morbidity of interest for all cancers combined and individual cancer sites. The numerator included those with any history of the relevant comorbidity at the given time point, except for obesity, which was classified based on the most recent body mass index (BMI) measure available at that time point.

#### Risk of influenza hospitalisation and mortality in cancer survivors and controls

Individuals were followed up from the index date until the earliest occurrence of the outcome, death without the outcome, or end of study period. Follow-up was not censored at the end of data collection in CPRD GOLD because the main analysis did not require post-baseline primary care data. We then fitted Cox proportional hazards models with time since index date as the timescale, initially accounting only for matching factors (i.e. age at index date, sex, and general practice) through stratification by matched set and then additionally adjusting for the presence of risk factors at the index date (for this analysis obesity was classified at the cancer diagnosis date since weight measures in the year following cancer diagnosis may be unstable). We examined the role of time since cancer diagnosis, by fitting a time-updated variable indicating time of cancer survivorship (1 to <5, 5 to <10, and ≥10 years since diagnosis, vs control group patient).

Since haematological malignancies directly affect the immune system and treatments may have long-term immune consequences, we stratified results by haematological versus other cancers by fitting a three-level cancer survivorship variable. Due to limited power, we did not break cancer sites down further. In a post hoc analysis, the exposure variable included each haematological malignancy separately (leukaemia, non-Hodgkin lymphoma, multiple myeloma) and four groups of solid cancers (i.e. breast, gastrointestinal, genitourinary, others); Wald tests were used after model estimation to test the null hypothesis of heterogeneity of effect among subgroups.

As a secondary analysis, we explored mediation of any raised risk of the primary outcome by development of recognised risk factors during follow-up, by adjusting for time-updated risk factor variables (taking the value “0” until the risk factor is first present, and “1” afterwards). This analysis was additionally censored at the end of follow-up in CPRD GOLD, since it relies on post-baseline primary care data.

Patients with missing data on BMI, smoking or deprivation were excluded from the cohorts (see above), therefore all models were based on complete case analyses. Multiple imputation was not used, as the missingness was considered likely to be not at random in the primary care setting ^15,16^, and complete case analysis minimises bias in this situation, providing missingness is conditionally independent of the outcome ^17^.*Sensitivity analyses:* We conducted two main sensitivity analyses. First, we broadened our definition of the outcome to include influenza recorded anywhere in the hospitalisation or death record, to account for the possibility of differential prioritisation of influenza codes between cancer survivors and controls. Second, we adjusted for time-updated influenza vaccination status and ever receipt of a pneumococcal vaccine, as cancer survivors may be more likely to receive influenza and pneumococcal vaccinations than general population controls due to higher engagement with healthcare, or vaccination indicated by immunosuppression following cancer and its treatment, which may protect against influenza and influenza-related death from secondary bacterial pneumonia. Influenza vaccinations were considered current from the date of vaccination until the start of the following influenza season in September. As vaccination records were ascertained from primary care data, these analyses were additionally censored at the end of CPRD follow up; we also re-ran the primary model with this additional censoring in order to provide a similarly censored comparator for the sensitivity and mediation analysis models.

#### Ethics

This study was approved by the London School of Hygiene & Tropical Medicine Ethics Committee (LSHTM Ethics Ref: 22416) and the Independent Scientific Advisory Committee for the Medicines and Healthcare products Regulatory Agency database research (20_082). Individual consent was not required for this study. CPRD supplies anonymised data for public health research; individuals are free to opt-out from having their data included in the database.

## Results

This study included 108,215 cancer survivors, of which 9,685 had prior haematological malignancies, and 523,541 individuals with no history of cancer (Table 1). Median (interquartile range [IQR]) age was 67 (58, 76) in the cancer survivor and comparison group; 6,674 (52.4%) and 277,781 (53.1%) of subjects were female, respectively.

### Prevalence of COVID-19 related risk factors in cancer survivors and controls

For all cancers combined, we observed higher absolute prevalence of all risk factors for severe COVID-19 except for obesity and sickle cell disease/splenectomy in 1-year cancer survivors, compared to the cancer-free comparison group (Figure 1, sickle cell/splenectomy not shown as the prevalence was <0.2% in all groups). At 5-years after diagnosis, cancer survivors overall had slightly higher prevalence of all risk factors except heart disease and neurological conditions (differences in prevalences ranging from 0.3% for diabetes and chronic liver disease, to 1.8% for chronic kidney disease). Survivors of most site-specific cancers also had raised prevalences of these risk factors, with the magnitude varying by cancer site. The prevalence of obesity was lower in cancer survivors than controls for several cancer sites, but was substantially more common in survivors of uterus and kidney cancers (prevalence difference at 5 years 20.1% [95%CI 19.8-20.5], and 8.5% [95%CI 8.1-8.8], respectively). Overall, 62.7% of the cancer survivors had at least 1 of the included comorbidities 5 years after diagnosis, while 37.3% had two or more (Supplementary Table S2). Comorbidity prevalences stratified by age and sex are provided in Supplementary Figure S2(a)-(h).

**Figure 1.**
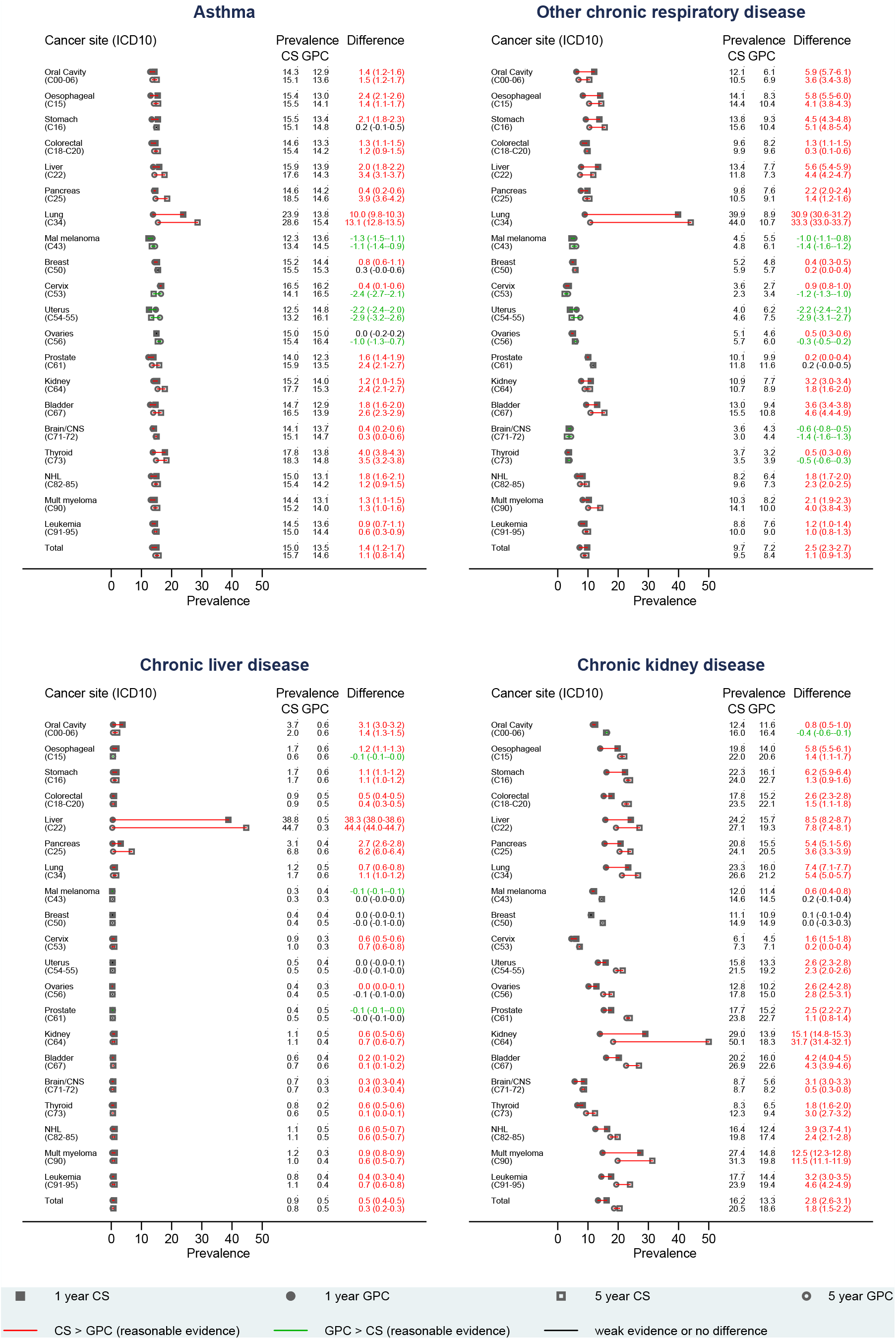
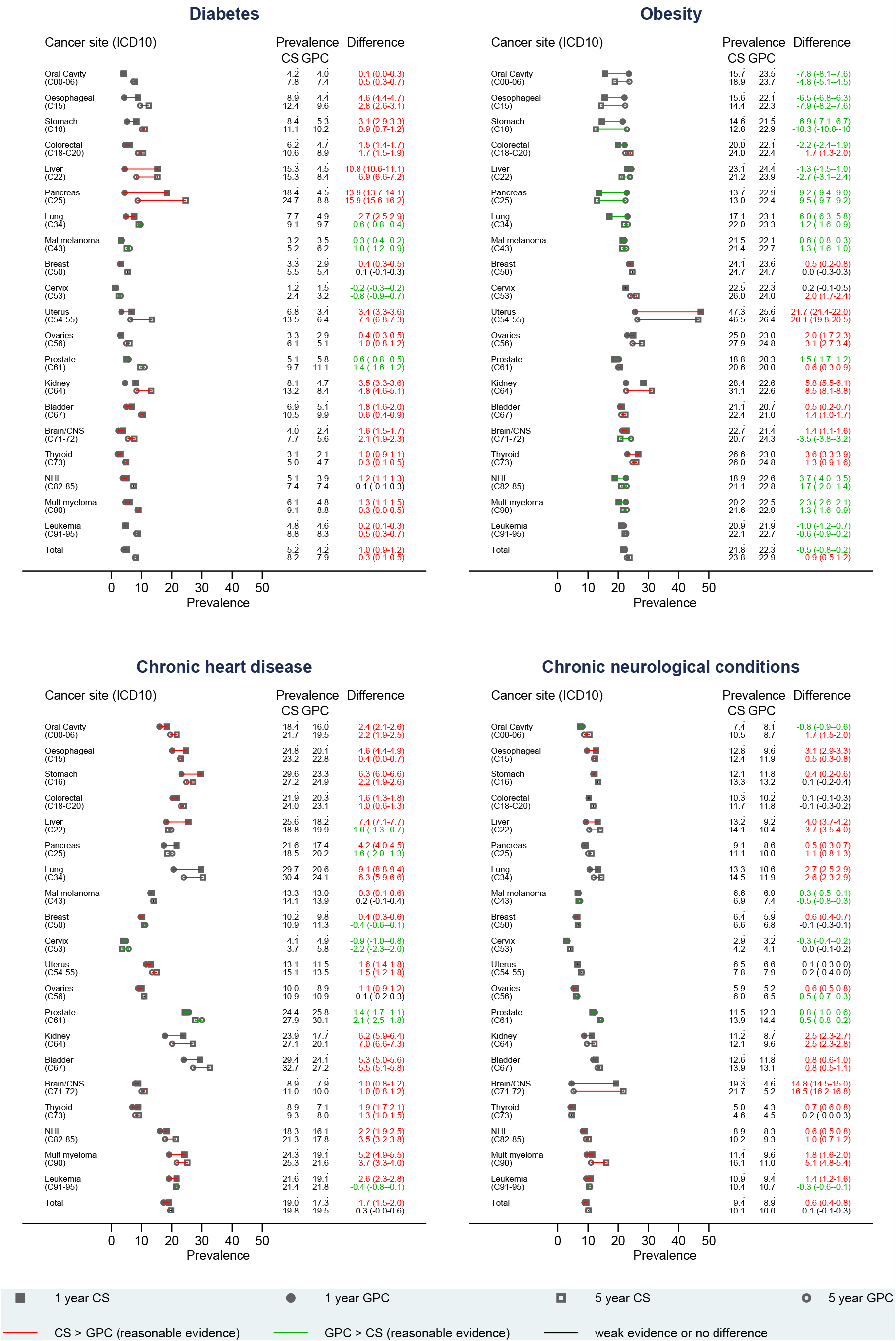
Prevalence of factors currently recognised as associated with high risk for severe COVID-19 outcomes in cancer survivors and controls at 1 and 5 years after diagnosis. Sickle cell disease and splenectomy are not presented due to the rarity of the outcome.

### Risk of influenza hospitalisation and mortality in cancer survivors and controls

205 people had the primary outcome (190 hospitalisations, 15 deaths) during a median follow-up time from the index date of 4.7 years in cancer survivors (IQR 1.9-8.4 years) and 6.2 years in controls (IQR 3.3-9.9 years); follow-up exceeded 10 years for 19,273 (18%) cancer survivors and 128,132 (25%) controls. The risk of influenza hospitalization or death was 2.7 times higher (95%CI 2.12-3.44) in cancer survivors compared to people with no history of cancer after accounting for matching factors only (Table 2). Control for other covariates had little impact on the relative risk estimate (adjusted HR=2.78; 95%CI 2.04-3.80).

**Table 2.** Relative risk of influenza hospitalisation or death in cancer survivors compared to non-cancer controls.

|  | No. of individuals | No. events<sup>#</sup> (no. deaths) | PY at risk | Associations adjusted for matching factors only* |  | Associations adjusted for risk factors for severe COVID-19 outcomes, smoking and IMD † |  |
| --- | --- | --- | --- | --- | --- | --- | --- |
|  |  |  |  | HR | 95% CI | HR | 95% CI |
| No cancer history | 523,541 | 205 (15) | 3,666,849 | Ref |  | Ref |  |
| All cancer survivors | 108,215 | 85 (6) | 620,144 | 2.70 | 2.12 – 3.44 | 2.78 | 2.04 – 3.80 |
| 1-5 years since diagnosis | 108,215 | 46 (2) | 326,913 | 4.16 | 2.94 – 5.88 | 4.34 | 2.86 – 6.59 |
| 5-10 years since diagnosis | 59,938 | 27 (3) | 201,332 | 2.15 | 1.44 – 3.21 | 2.37 | 1.53 – 3.66 |
| >10 years since diagnosis | 24,111 | 12 (1) | 91,899 | 1.38 | 0.74 – 2.56 | 1.37 | 0.74 – 2.52 |
| By cancer group |  |  |  |  |  |  |  |
| Haematological malignancies | 9,685 | 40 (3) | 52,573 | 12.94 | 7.47 – 22.44 | 15.17 | 7.84 – 29.35 |
| 1-5 years since diagnosis | 9,685 | 20 (1) | 29,072 | 22.21 | 8.34 – 59.17 | 29.56 | 10.20 – 85.66 |
| 5-10 years since diagnosis | 5,131 | 15 (2) | 16,608 | 8.62 | 4.00 – 18.58 | 9.56 | 4.39 – 20.84 |
| >10 years since diagnosis | 1,870 | ≤5 (0) | 6,894 | 9.90 | 2.46 – 39.78 | 10.06 | 2.47 – 40.93 |
| All other cancers | 98,530 | 45 (3) | 567,570 | 1.50 | 1.09 – 2.05 | 1.38 | 0.92 – 2.07 |
| 1-5 years since diagnosis | 98,530 | 26 (1) | 297,841 | 2.47 | 1.61 – 3.80 | 2.22 | 1.31 – 3.74 |
| 5-10 years since diagnosis | 54,807 | 12 (1) | 184,724 | 1.05 | 0.62 – 1.80 | 1.08 | 0.58 – 2.01 |
| >10 years since diagnosis | 22,241 | 7 (1) | 85,005 | 0.77 | 0.34 – 1.73 | 0.74 | 0.34 – 1.61 |

<sup>#</sup> Hospitalisations or deaths with influenza as the primary diagnosis/underlying cause.
\* Cancer survivors and non-cancer controls were matched on age (within a 3-year age range), sex and primary care practice.
† Model adjusted for risk factors for poor COVID-19 outcomes (i.e. asthma and other chronic respiratory diseases, chronic neurological diseases, chronic liver disease, chronic heart disease, chronic kidney disease, sickle cell disease or splenectomy, diabetes and obesity), smoking (former vs. current vs. never smokers), and quintiles of relative deprivation measured by patient-postcode linked Index of Multiple Deprivation.
HR = Hazards ratio; PY = person-years at risk; Ref = reference category.

Stratification by cancer group (haematological vs non-haematological) showed substantial differences. Haematological cancer survivors had 15 times higher risk of a severe influenza outcome compared to people without cancer (adjusted HR 15.17; 95%CI 7.84-29.35), and further stratifying by time since cancer diagnosis, the hazard ratio was 29.56 (95%CI 10.20-85.66) for those 1 to <5 years from diagnosis, falling to 9.56 (95%CI 4.39-20.84) and 10.06 (95%CI 2.47-40.93) for those 5 to <10, and 10+ years from diagnosis respectively. Associations were smaller for non-haematological cancer survivors. The overall adjusted HR was 1.38 but compatible with chance variation (95%CI 0.92-2.07). However, stratification by time since diagnosis suggested a doubling of risk in those 1 to <5 years from diagnosis (adjusted HR 2.22, 1.31-3.74) with no raised risk in longer-term survivors.

### Sensitivity, mediation and post-hoc analyses

Using hospitalisations and deaths with any mention of influenza in the outcome definition led to more events being included (n=320) but a very similar pattern of results to the primary analysis (Supplementary Table S3). In analyses that censored at end of CPRD follow-up, fewer events were included (n=167) but hazard ratios were generally larger than in the primary analysis (overall adjusted HR for cancer survivors vs controls 3.88, 2.54-5.91, Supplementary Table S4). Additional control for time-updated exposure to influenza and pneumococcal vaccination led to similar but slightly stronger associations (overall HR 4.06, 2.65-6.24), while adjusting for mediators led to slightly weaker associations (overall HR 3.27, 95%CI 2.12-5.04), but in both cases patterns of results were similar. There was no strong statistical evidence of a variation in the HRs among survivors of leukaemia, non-Hodgkin lymphoma and multiple myeloma (p=0.08), or among survivors from the different solid cancers (p=0.42).

## Discussion

Most comorbidities thought to be risk factors for COVID-19 were more prevalent in cancer survivors than cancer-free controls, with variation by cancer site. After accounting for baseline demographics, deprivation, smoking and risk factors distribution, the risks of influenza hospitalisation and death were elevated >9-fold in haematological cancer survivors compared with matched controls for at least 10 years after diagnosis, and >2-fold in non-haematological cancer survivors in the one to five years after diagnosis.

To our knowledge, this is the first large cohort study using prospectively collected data to quantify the relative risk of severe influenza outcomes in different groups of cancer survivors compared to the general population, including stratification by time since diagnosis. The few previous studies in this area have reported high rates of influenza among cancer survivors, consistent with our findings, but have lacked a cancer-free comparison group.^18-21^ Hermann et al. investigated outcomes among patients with a history of cancer presenting with influenza, and found no difference in mortality according to haematological or non-haematological cancer type, or activity of the cancer.^18^ Our results showed considerably higher risks of hospitalisation or death among haematological cancer survivors, which could be consistent with the findings in Hermann et al. if haematological cancer survivors are at increased risk of infection, but not mortality once infected, compared to non-haematological cancer survivors. Other studies have investigated vulnerability to influenza infection of any severity; two studies using administrative claims data in South Korea found a high rate of claims for influenza among both breast cancer survivors and survivors of childhood cancers.^19,20^ Similarly, Australian survey data found that a large proportion (38%) of hematopoietic stem cell transplant survivors had had influenza-like illnesses in the time (median 5 years) since their transplant suggesting potentially high vulnerability to infection, but there was no control group or information on severity of infection.^21^

Direct evidence on how COVID-19 affects cancer patients and survivors is immature. Early evidence from China and Italy suggested that patients with history of cancer were overrepresented among those admitted to hospital with COVID-19.^22,23^ The large UK OpenSAFELY study found substantially raised risks of COVID-19 mortality among individuals with prior haematological cancer persisting for at least 5 years from cancer diagnosis, and smaller raised risks for those with a history of non-haematological cancers up to 5 years from diagnosis, consistent with our findings for influenza.^11^ A study from the COVID-19 and Cancer Consortium (CCC19) reported high 30-day mortality among individuals with laboratory-confirmed COVID-19 and active or previous malignancy, finding high 30-day mortality, even among those in remission, though active disease was a strong predictor of mortality.^24^ Finally, a study that focussed on patients with active cancer and COVID-19 found a non-statistically significant increased risk of mortality in patients exposed to chemotherapy 4 weeks prior to infection (OR=1.18, 95%CI 0.81-1.72), compared to cancer patients that did not receive chemotherapy, but the small numbers involved require further studies to confirm these associations.^25^

We used a large cohort of cancer survivors and matched controls, nearly a quarter of whom were followed up for more than 10 years. The size of our study enabled us to estimate prevalence of risk factors for severe respiratory infections in site-specific cancer survivors for the twenty most common cancer sites with good precision, and to adjust our primary analysis of severe influenza outcomes for multiple risk factors and stratify by type of cancer (haematological vs other). Multiple validation studies have demonstrated the validity of CPRD primary care data for measuring disease phenotypes including cancer, especially when combined with additional linked data sources.^26^ Our primary analysis was designed to be specific to hospitalisations and deaths caused by influenza, and a broader definition in sensitivity analysis found similar results. A second sensitivity analysis took account of time-updated vaccination status, which showed that the associations we observed persisted, and in fact were stronger after accounting for this apparent negative confounder.

There are some important limitations. We analysed severe influenza in an attempt to inform COVID-19 policy but despite both being infectious respiratory illnesses, it is not certain that risk factors for severe influenza will have the same associations with COVID-19. Our approach follows that of policy makers who have assumed parallels with influenza in the absence of mature COVID-19 data.^27^ As data from the COVID-19 pandemic itself have started to flow, they have largely confirmed a broad overlap between those at high risk for seasonal influenza and for severe COVID-19 outcomes.^11^ Another limitation was that we did not have data on anti-cancer treatments, so could not separate cancer survivors into those under active treatment or not undergoing any treatment, which may be an important determinant of risk. We only included cancer survivors at least one year out from diagnosis, so it is likely that most patients with high-grade malignancies would have completed primary treatment, but people with low-grade tumours could conceivably have received anticancer therapies some years after initial diagnosis, which could explain part of the medium- to long-term increased risk of severe influenza; linked cancer treatment data will be needed to investigate this further. We cannot rule out that differences in the prevalence of risk factors between cancer survivors and controls five years post-diagnosis may be due to increased contact with health services, particularly for diseases such as chronic kidney disease which may be asymptomatic. Our primary outcome combined influenza hospitalisations and deaths but was dominated by the former; it is plausible that there may be a lower threshold for hospitalisation in cancer survivors which could have exaggerated the difference in risk of the primary outcome between cancer survivors and controls, but is unlikely to fully explain the large associations we observed. Finally, we had some missing data on smoking and BMI data, and we excluded those with missing data from the analysis; this is unlikely to affect our findings under the assumption that the association between cancer survivorship and severe respiratory outcomes is the same in people with and without missing data, conditional on the covariates included in the model. We have no reason to doubt this assumption, as recording of BMI and smoking in primary care could be associated with cancer survivorship but most likely is not associated with the risk of influenza hospitalization or death.

The high prevalence of several established COVID-19 risk factors in cancer survivors, and the increased risk of influenza hospitalisation and death in survivors of haematological cancers even many years from diagnosis, and in survivors from other cancers in the first five years of survivorship, indicate a likely increased risk of severe COVID-19 outcomes in these patient groups. Early direct evidence from the COVID-19 pandemic appears to be consistent with this. These findings suggest that cancer survivorship should be considered a potentially important risk factor for severe COVID-19 outcomes in public health policy. At present, while UK policy defines those with active cancers and/or receiving treatments as high-risk for COVID-19 complications, the much larger overall population of cancer survivors does not appear in either moderate or high-risk groupings;^28^ these risk groupings become increasingly important as general population social distancing measures are eased and advice becomes more targeted to those at risk.

Our results also have implications for preventive medicine in the coming autumn and winter, when influenza and SARS-CoV-2 are expected to coexist in the population. Improving influenza vaccination coverage among cancer survivors should be a priority, as the vaccine is both effective and safe^29,30^ but coverage has been reported in the range of 50% to 76% among cancer survivors in the US and in the UK.^31,32^ Immunisation for *streptococcus pneumoniae* may also be considered.^33^ Of note, UK influenza vaccine guidance focusses on cancer patients with active or recent disease or treatment;^5^ our findings suggest that a broader population of cancer survivors should be considered as a high-risk group for influenza vaccination.

Future studies should focus on the risk of severe COVID-19 in cancer survivors, explore the role of comorbidities and prior exposure to specific anti-cancer therapies, disaggregating data by cancer site when possible.

In conclusion, survivors of haematological malignancies had substantially elevated risks of influenza hospitalisation or death persisting for at least 10 years after cancer diagnosis, while risk was doubled for survivors of other cancers for up to 5 years from diagnosis. In addition, cancer survivors had higher prevalence of several chronic conditions associated with severe COVID-19, compared to people with no history of cancer. This should be taken into account in public health policies targeted at protecting clinical risk groups. Influenza vaccination should be encouraged in this group, and may need to be extended to a wider population of medium- to long-term cancer survivors than currently recommended.

## Data Availability

This study is based in part on data from the Clinical Practice Research Datalink obtained under licence from the UK Medicines and Healthcare products Regulatory Agency. The terms of our licence to access the data preclude us from sharing individual patient data with third parties. The raw data may be requested directly from CPRD following their usual procedures.

## Acknowledgements

This study is based in part on data from the Clinical Practice Research Datalink obtained under licence from the UK Medicines and Healthcare products Regulatory Agency. The data is provided by patients and collected by the NHS as part of their care and support. The interpretation and conclusions contained in this study are those of the author/s alone. The study was approved by the Independent Scientific Advisory Committee (approval number: 20_082).

## Contributors

KB, HS and HC designed the study. HS and KB created the data set for a previous study. MP and HMcD created code lists to identify immunisations in the primary care data. HC, HS and KB conducted the analyses in the present study. HC and HS wrote the first draft of the manuscript. All authors revised the manuscript for important intellectual content. HC, HS and KB are guarantors for this study, had access to all study data and accept full responsibility for the work.

## Supplementary Materials

Supplementary table S1. Definition of the study variables.

Supplementary table S2. Number of comorbidities at 1 and 5 years after cancer diagnosis.

Supplementary table S3. Results from sensitivity analysis additionally including outcomes where influenza was present but not considered the primary diagnosis in the hospitalization, and/or the primary cause of death.

Supplementary table S4. Results from sensitivity and mediation analyses showing the relative risk of influenza hospitalisation or death in cancer survivors compared to cancer-free controls.

Supplementary figure S1. Flow chart of people included in the study.

Supplementary figure S2. Prevalence of factors currently recognised as associated with high risk for severe COVID-19 outcomes in cancer survivors and controls stratified by age group and sex.

