## Supplementary for "Prevalence of COVID-19-related risk factors and risk of severe influenza outcomes in cancer survivors: a matched cohort study using linked English electronic health records data"

**Supplementary table S1**. Definition of the study variables.

| **Variable** | **Definition** | **List of odes** |
| --- | --- | --- |
| Asthma | Any previous diagnosis of asthma. | Read codes from Nissen et al. [BMJ Open. 2017 Aug 11;7(8):e017474](https://www.ncbi.nlm.nih.gov/pmc/articles/PMC5724126/) |
| Chronic respiratory diseases other than asthma | Any previous diagnosis of a long-term respiratory disease, such as chronic obstructive pulmonary disease. | Read codes available at: <https://doi.org/10.17037/DATA.00001663> |
| Chronic heart disease | Any diagnosis of chronic heart disease likely to need follow up or medication. | Read codes available at: <https://doi.org/10.17037/DATA.00001661> |
| Chronic liver disease | Any previous diagnosis of a chronic liver disease including cirrhosis, oesophageal varices, biliary atresia and chronic hepatitis. | Read codes available at: <https://doi.org/10.17037/DATA.00001662> |
| Chronic neurological disease | Conditions in which respiratory function may be compromised due to chronic neurological disease, including stroke, transient ischaemic attack, cerebral palsy, Parkinson's disease and multiple sclerosis. | Read codes available at: <https://doi.org/10.17037/DATA.00001704> |
| Chronic kidney disease (CKD)  (based on diagnoses and serum creatinine test results) | Any previous history of dialysis or renal transplant; or a Read code recording CKD stage 3-5; or nephrotic syndrome without a more recent Read code recording CKD stage 1-2; or latest estimated glomerular filtration rate (eGFR) ≤60mL/min/1.73m^2^ using serum creatinine test results. | Read codes available at: <https://doi.org/10.17037/DATA.00001664> |
| Diabetes mellitus | Any previous diagnosis of diabetes mellitus, or a diagnosis within the previous three months if a reversible cause was specified. | Read codes available at: <https://doi.org/10.17037/DATA.00001665> |
| Obesity | Body mass index ≥30 kg/m^2^, calculated from weight and height. | Algorithm from Bhaskaran et al. [BMJ Open. 2013; 3(9): e003389](https://www.ncbi.nlm.nih.gov/pmc/articles/PMC3773634/) |
| Sickle cell disease/ splenectomy | Any previous diagnosis of sickle cell disease or having undergone splenectomy. | Read codes from Grint et al. [BMJ Open 2020 Jan 29;10(1):e034886](https://bmjopen.bmj.com/content/10/1/e034886.long). |
| Smoking | Patients classified as current smokers, former smokers and never smokers at study baseline. | Read codes available at: <https://doi.org/10.17037/DATA.00001602> |
| Index of Multiple Deprivation | Patient-postcode linked quintiles of relative deprivation. | Official measure of relative deprivation for small areas in England obtained through data linkage. |
| Influenza vaccination | Immunisation with seasonal and/or pandemic influenza vaccine. Considered immunized from the date of the vaccine record up to the beginning of the next year season (seasons from 1^st^ of September up to August 31^st^ of the following year). | Read codes available at: <https://doi.org/10.17037/DATA.00001626>;  <https://doi.org/10.17037/DATA.00001627>;  <https://doi.org/10.17037/DATA.00001628>. |
| Pneumococcal vaccination | Any record of having received a vaccine for *streptococcus* *pneumoniae*. | Read codes available at: <https://doi.org/10.17037/DATA.00001808>. |
| Influenza hospitalization | Primary diagnosis of influenza infection in the Hospital Episode Statistics dataset. | ICD-10 codes J09-J11. |
| Death due to influenza | Deaths registered in the mortality data from the Office for National Statistics with influenza as the main cause. | ICD-10 codes J09-J11;  ICD-9 codes 487-488. |

**Supplementary table S2**. Number of comorbidities at 1 and 5 years after cancer diagnosis.

|  |  | **1 year after diagnosis** | | |  | | **5 years after diagnosis** | |
| --- | --- | --- | --- | --- | --- | --- | --- | --- |
|  |  | **≥1**  **comorbidity** | **≥2 comorbidities** |  | | **≥1 comorbidity** | | **≥2 comorbidities** |
|  |  | **N (%)** | **N (%)** |  | | **N (%)** | | **N (%)** |
| **Bladder** | CS | 4,985 (64.6) | 2,590 (33.6) |  | | 5,959 (77.3) | | 3,770 (48.9) |
|  | GPC | 21,687 (58.8) | 9,957 (27.0) |  | | 25,523 (69.2) | | 14,815 (40.2) |
| **Breast** | CS | 12,420 (48.5) | 4,730 (18.5) |  | | 14,697 (57.3) | | 6,863 (26.8) |
|  | GPC | 59,815 (47.4) | 21,692 (17.2) |  | | 70,110 (55.6) | | 32,064 (25.4) |
| **Cervix** | CS | 489 (40.4) | 154 (12.7) |  | | 589 (48.7) | | 208 (17.2) |
|  | GPC | 2,448 (40.0) | 673 (11.0) |  | | 2,815 (46.0) | | 965 (15.8) |
| **Brain/CNS** | CS | 472 (52.1) | 186 (20.5) |  | | 549 (60.6) | | 253 (27.9) |
|  | GPC | 1,808 (41.4) | 556 (12.7) |  | | 2,102 (48.2) | | 824 (18.9) |
| **Colorectal** | CS | 8,415 (59.2) | 3,943 (27.7) |  | | 9,928 (69.8) | | 5,693 (40.0) |
|  | GPC | 38,896 (56.6) | 17,089 (24.8) |  | | 45,893 (66.7) | | 25,496 (37.1) |
| **Stomach** | CS | 967 (64.2) | 527 (35.0) |  | | 1,067 (70.8) | | 648 (43.0) |
|  | GPC | 4,172 (58.5) | 1,940 (27.2) |  | | 4,931 (69.1) | | 2,896 (40.6) |
| **Kidney** | CS | 1,505 (68.5) | 779 (35.5) |  | | 1,834 (83.5) | | 1,157 (52.7) |
|  | GPC | 5,645 (54.1) | 2,481 (23.8) |  | | 6,596 (63.2) | | 3,505 (33.6) |
| **Leukaemia** | CS | 2,031 (59.4) | 928 (27.1) |  | | 2,425 (70.9) | | 1,377 (40.3) |
|  | GPC | 8,964 (54.5) | 3,928 (23.9) |  | | 10,582 (64.3) | | 5,789 (35.2) |
| **Liver** | CS | 431 (77.8) | 274 (49.5) |  | | 467 (84.3) | | 331 (59.7) |
|  | GPC | 1,490 (56.4) | 650 (24.6) |  | | 1,729 (65.4) | | 906 (34.3) |
| **Lung** | CS | 4,064 (75.7) | 2,485 (46.3) |  | | 4,426 (82.4) | | 3,056 (56.9) |
|  | GPC | 15,195 (58.2) | 6,910 (26.5) |  | | 17,688 (67.7) | | 9,903 (37.9) |
| **Malignant melanoma** | CS | 3,355 (47.3) | 1,218 (17.2) |  | | 3,943 (55.6) | | 1,838 (25.9) |
|  | GPC | 16,446 (48.2) | 6,329 (18.6) |  | | 19,200 (56.3) | | 9,123 (26.7) |
| **Multiple myeloma** | CS | 1,213 (65.8) | 598 (32.4) |  | | 1,463 (79.4) | | 899 (48.8) |
|  | GPC | 4,881 (55.6) | 2,163 (24.6) |  | | 5,842 (66.5) | | 3,189 (36.3) |
| **Non-Hodgkin lymphoma** | CS | 2,478 (56.0) | 1,071 (24.2) |  | | 2,931 (66.3) | | 1,581 (35.7) |
|  | GPC | 11,011 (52.0) | 4,432 (20.9) |  | | 12,946 (61.1) | | 6,560 (31.0) |
| **Oesophageal** | CS | 1,170 (65.2) | 607 (33.8) |  | | 1,249 (69.6) | | 726 (40.5) |
|  | GPC | 4,730 (55.3) | 2,133 (24.9) |  | | 5,621 (65.7) | | 3,091 (36.1) |
| **Oral Cavity** | CS | 865 (54.6) | 371 (23.4) |  | | 988 (62.4) | | 512 (32.3) |
|  | GPC | 4,024 (51.7) | 1,607 (20.6) |  | | 4,693 (60.3) | | 2,368 (30.4) |
| **Ovaries** | CS | 1,387 (51.2) | 497 (18.3) |  | | 1,672 (61.7) | | 753 (27.8) |
|  | GPC | 6,234 (46.3) | 2,211 (16.4) |  | | 7,364 (54.7) | | 3,271 (24.3) |
| **Pancreas** | CS | 549 (63.5) | 277 (32.1) |  | | 637 (73.7) | | 366 (42.4) |
|  | GPC | 2,257 (55.2) | 953 (23.3) |  | | 2,639 (64.5) | | 1,360 (33.3) |
| **Prostate** | CS | 12,382 (59.8) | 5,666 (27.4) |  | | 14,817 (71.5) | | 8,656 (41.8) |
|  | GPC | 58,724 (59.5) | 27,314 (27.7) |  | | 69,200 (70.1) | | 40,320 (40.9) |
| **Thyroid** | CS | 492 (47.9) | 184 (17.9) |  | | 554 (53.9) | | 253 (24.6) |
|  | GPC | 2,073 (42.0) | 612 (12.4) |  | | 2,430 (49.3) | | 884 (17.9) |
| **Uterus** | CS | 2,256 (65.6) | 949 (27.6) |  | | 2,510 (73.0) | | 1,390 (40.4) |
|  | GPC | 8,709 (51.5) | 3,406 (20.1) |  | | 10,234 (60.5) | | 4,969 (29.4) |
| **Total** | CS | 61,926 (57.2) | 28,034 (25.9) |  | | 72,705 (67.2) | | 40,330 (37.3) |
|  | GPC | 279,209 (53.3) | 117,036 (22.4) |  | | 328,138 (62.7) | | 172,298 (32.9) |

CS = Cancer Survivors; GPC = General Population Control

**Supplementary table S3.** Results from sensitivity analysis additionally including outcomes where influenza was present but not considered the primary diagnosis in the hospitalization, and/or the primary cause of death.

|  |  | | |  | **Associations adjusted for matching factors only*** | |  | **Associations adjusted for risk factors for severe COVID-19 outcomes, smoking and IMD** † | |
| --- | --- | --- | --- | --- | --- | --- | --- | --- | --- |
|  | **No. of individuals** | **No. of events^#^**  **(no. deaths)** | **PY at risk** |  | **HR** | **95% CI** |  | **HR** | **95% CI** |
| **All cancer survivors** |  |  |  |  |  |  |  |  |  |
| No cancer history | 523,541 | 320 (11) | 3,666,555 |  | Ref |  |  | Ref |  |
| Cancer survivors | 108,215 | 142 (7) | 620,024 |  | 3.01 | 2.50 – 3.64 |  | 2.96 | 2.32 – 3.77 |
| 1-4 years | 108,215 | 86 (3) | 326,872 |  | 4.41 | 3.41 – 5.70 |  | 4.35 | 3.19 – 5.91 |
| 5-9 years | 59,924 | 37 (3) | 201,285 |  | 2.01 | 1.43 – 2.82 |  | 2.10 | 1.44 – 3.06 |
| ≥10 years | 24,105 | 19 (1) | 91,867 |  | 1.86 | 1.13 – 3.04 |  | 1.75 | 1.01 – 3.02 |
| **By cancer group** |  |  |  |  |  |  |  |  |  |
| Haematological malignancies | 9,685 | 64 (4) | 52,532 |  | 12.38 | 8.02 – 19.10 |  | 15.11 | 8.95 – 25.53 |
| 1-4 years | 9,685 | 38 (2) | 29,053 |  | 26.87 | 12.12 – 59.56 |  | 34.95 | 15.54 – 78.57 |
| 5-9 years | 5,125 | 18 (2) | 16,595 |  | 6.32 | 3.35 – 11.91 |  | 7.62 | 3.96 – 14.67 |
| ≥10 years | 1,869 | 8 (0) | 6,884 |  | 7.76 | 2.84 – 21.23 |  | 8.48 | 2.90 – 24.79 |
| All other cancers | 98,530 | 78 (3) | 567,492 |  | 1.80 | 1.42 – 2.29 |  | 1.55 | 1.14 – 2.12 |
| 1-4 years | 98,530 | 48 (1) | 297,819 |  | 2.62 | 1.92 – 3.59 |  | 2.21 | 1.49 – 3.26 |
| 5-9 years | 54,799 | 19 (1) | 184,690 |  | 1.18 | 0.76 – 1.83 |  | 1.11 | 0.66 – 1.86 |
| ≥10 years | 22,236 | 11 (1) | 84,983 |  | 1.12 | 0.59 – 2.12 |  | 0.98 | 0.49 – 1.95 |

^#^ Hospitalisations or deaths with influenza mentioned anywhere in the HES record/death certificate.

***** Cancer survivors and non-cancer controls were matched on age (within a 3-year age range), sex and primary care practice.

† The model was also adjusted for risk factors for poor COVID-19 outcome (i.e. asthma and other chronic respiratory diseases, chronic neurological diseases, chronic liver disease, chronic heart disease, chronic kidney disease, sickle cell disease or splenectomy, diabetes and obesity), smoking (former vs. current vs. never smokers), and quintiles of relative deprivation measured by patient-postcode linked Index of Multiple Deprivation.

HR = Hazards ratio; IMD = Index of Multiple Deprivation; PY = person-years at risk; Ref = reference category.

**Supplementary table S4**. Results from sensitivity and mediation analyses showing the relative risk of influenza hospitalisation or death in cancer survivors compared to cancer-free controls.

|  |  |  |  |  | **Models adjusted for risk factors for severe COVID-19 outcomes, smoking and IMD, censored at end of CPRD follow up** | |  | **Models adjusted for influenza vaccination*** | |  | **Models adjusted for influenza & pneumococcal vaccination** † | |  | **Mediation analysis** | |
| --- | --- | --- | --- | --- | --- | --- | --- | --- | --- | --- | --- | --- | --- | --- | --- |
|  | **No. of individuals** | **No. of events^#^ (no. deaths)** | **PY at risk** |  | **HR** | **95% CI** |  | **HR** | **95% CI** |  | **HR** | **95% CI** |  | **HR** | **95% CI** |
| No cancer history | 523,541 | 110 (7) | 2,817,699 |  | Ref |  |  | Ref |  |  | Ref |  |  | Ref |  |
| All cancer survivors | 108,215 | 57 (1) | 480,766 |  | 3.88 | 2.54-5.91 |  | 4.06 | 2.65-6.22 |  | 4.08 | 2.63-6.34 |  | 3.27 | 2.12-5.04 |
| 1-5 years since diagnosis | 108,215 | 36 (0) | 289,392 |  | 5.11 | 3.07-8.52 |  | 5.48 | 3.24-9.27 |  | 5.46 | 3.23-9.24 |  | - | - |
| 5-9 years since diagnosis | 46,822 | 17 (1) | 143,038 |  | 2.95 | 1.67-5.23 |  | 3.01 | 1.60-5.66 |  | 3.03 | 1.62-5.70 |  | - | - |
| ≥10 years since diagnosis | 14,995 | ≤5 (0) | 48,336 |  | 1.67 | 0.62-4.53 |  | 1.79 | 0.63-5.09 |  | 1.79 | 0.63-5.03 |  | - | - |
| **By cancer group** |  |  |  |  |  |  |  |  |  |  |  |  |  |  |  |
| Haematological malignancies | 9,685 | 30 (1) | 40,527 |  | 26.90 | 10.18-71.06 |  | 38.05 | 13.74-105.36 |  | 38.16 | 13.71-106.21 |  | 19.22 | 7.55-48.93 |
| 1-4 years since diagnosis | 9,685 | 16 (0) | 25,767 |  | 39.57 | 9.41-166.38 |  | 58.35 | 13.80-246.77 |  | 59.11 | 13.43-260.19 |  | - | - |
| 5-9 years since diagnosis | 4023 | 12 (1) | 11,528 |  | 15.98 | 4.99-51.18 |  | 22.74 | 5.60-92.31 |  | 22.74 | 5.58-92.70 |  | - | - |
| ≥10 years since diagnosis | 1098 | ≤2 (0) | 3,233 |  | - | - |  | - | - |  | - | - |  | - | - |
| All other cancers | 98,530 | 27 (0) | 440,239 |  | 1.65 | 0.94-2.88 |  | 1.59 | 0.90-2.80 |  | 1.59 | 0.90-2.80 |  | 1.41 | 0.79-2.52 |
| 1-4 years since diagnosis | 98,530 | 20 (0) | 263,625 |  | 2.60 | 1.45-4.67 |  | 2.65 | 1.45-4.83 |  | 2.64 | 1.45-4.82 |  | - | - |
| 5-9 years since diagnosis | 42,799 | ≤5 (0) | 131,511 |  | 0.71 | 0.29-1.76 |  | 0.59 | 0.22-1.61 |  | 0.59 | 0.122-1.61 |  | - | - |
| ≥10 years since diagnosis | 13,897 | ≤2 (0) | 45,103 |  | 0.60 | 0.15-2.32 |  | 0.64 | 0.09-4.58 |  | 0.63 | 0.09-4.60 |  | - | - |
| h |  |  |  |  |  |  |  |  |  |  |  |  |  |  |  |

^#^ Hospitalisations or deaths with influenza as the primary diagnosis/underlying cause.

* Influenza vaccination was added to the model as a time updated covariate. We considered that a patient was immunized from the date of the vaccine record until the day prior to the beginning of the following flu season. The model was also adjusted for risk factors for poor COVID-19 outcome (i.e. asthma and other chronic respiratory diseases, chronic neurological diseases, chronic liver disease, chronic heart disease, chronic kidney disease, sickle cell disease or splenectomy, diabetes and obesity), smoking (former vs. current vs. never smokers), and quintiles of relative deprivation measured by patient-postcode linked Index of Multiple Deprivation. Cancer survivors and non-cancer controls were matched on age (within a 3-year age range), sex and primary care practice.

† Pneumococcal vaccination defined as a binary variable (ever/never).HR = Hazards ratio; PY = person-years at risk; Ref = reference category.

**Supplementary Figure S1.** Flow chart of people included in the study.

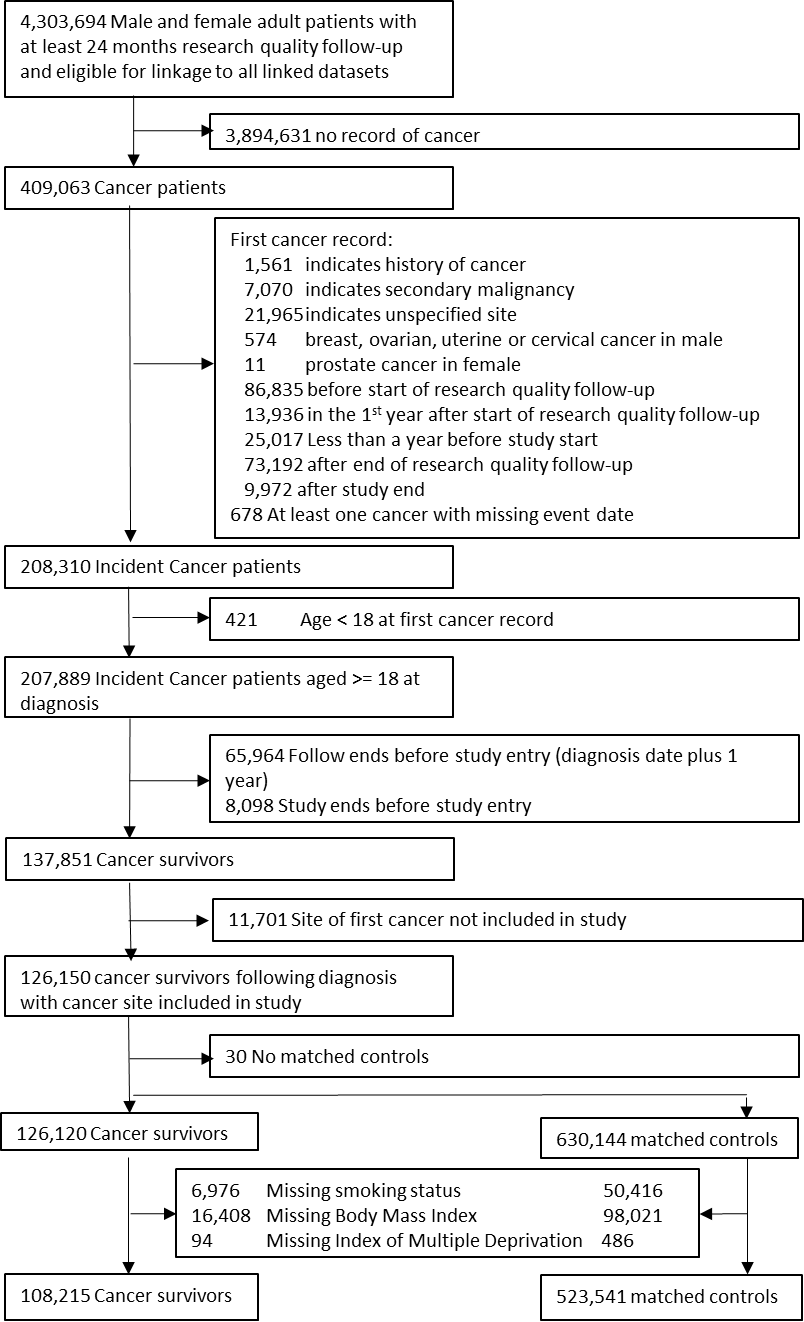

Supplementary figure S2. Prevalence of factors currently recognised as associated with high risk for severe COVID-19 outcomes in cancer survivors and controls stratified by age group and sex.

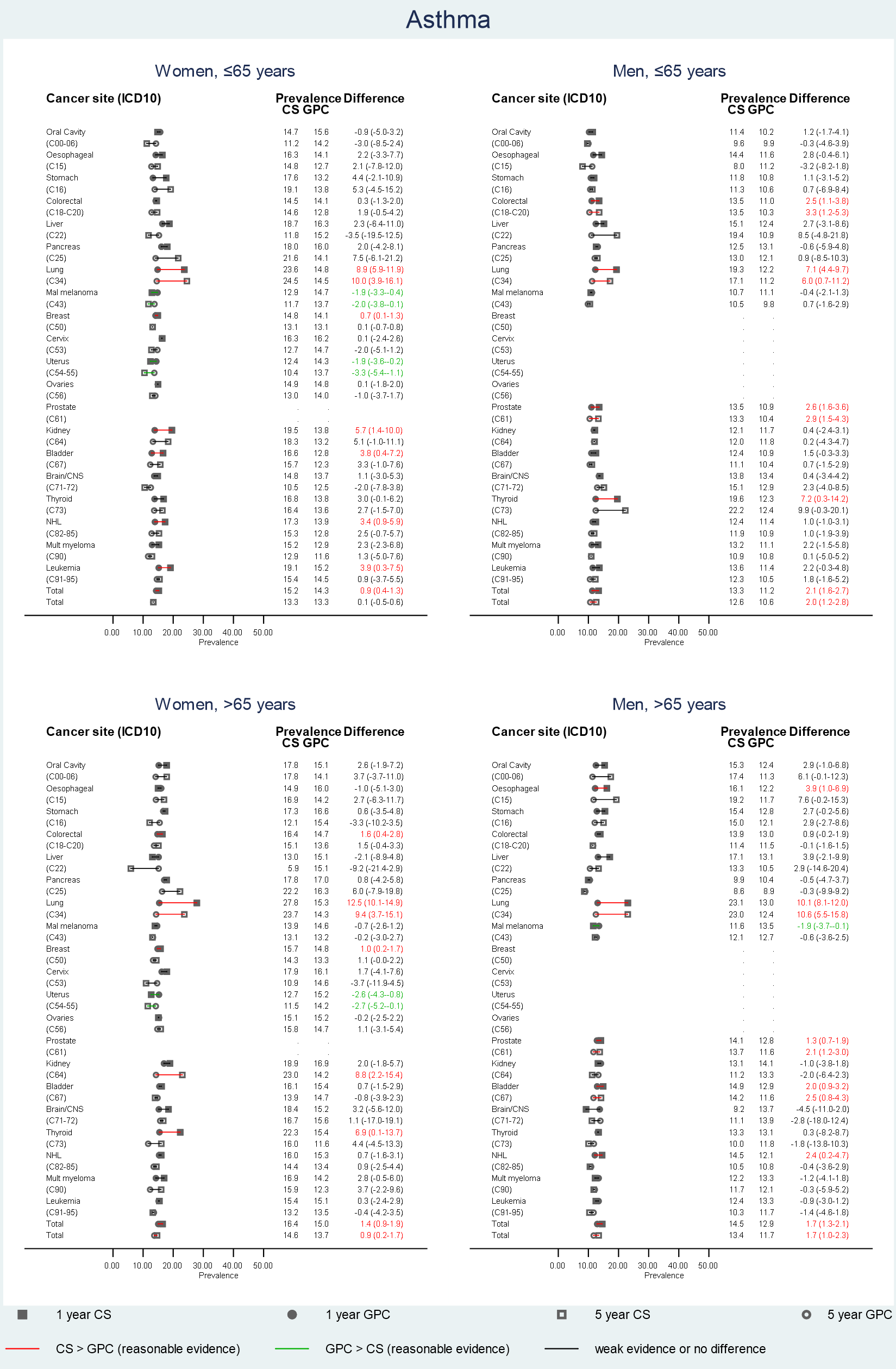

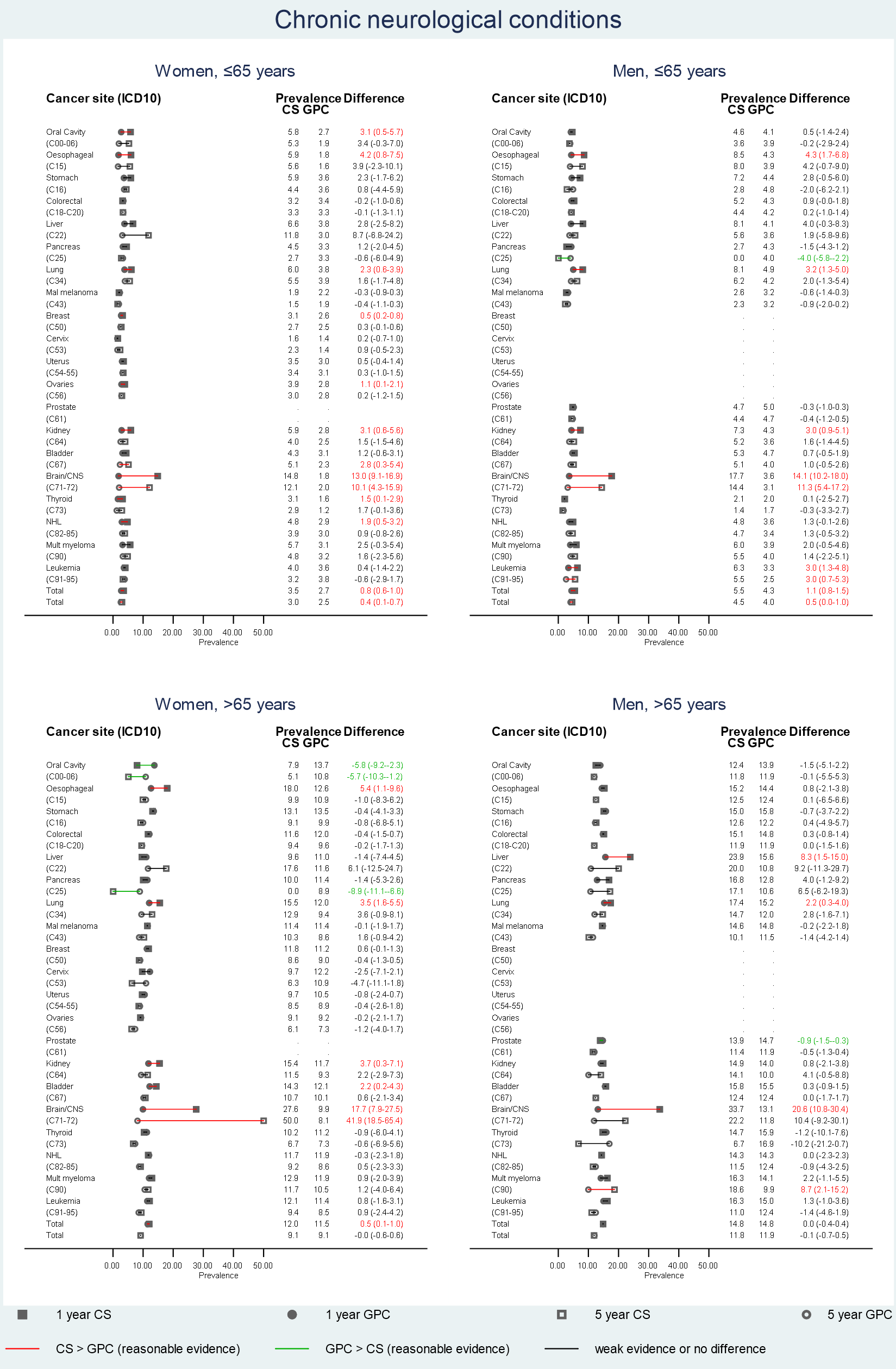

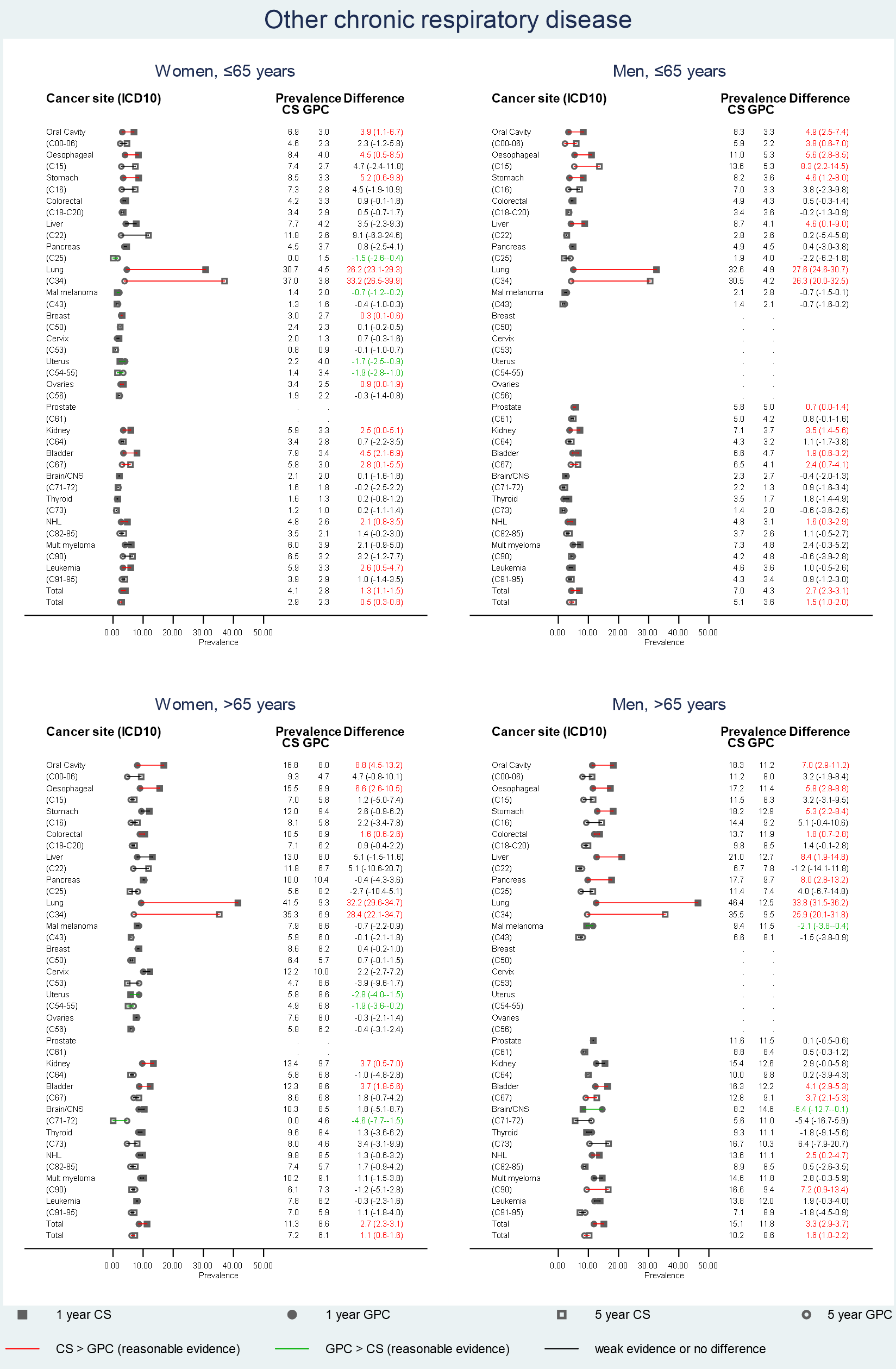

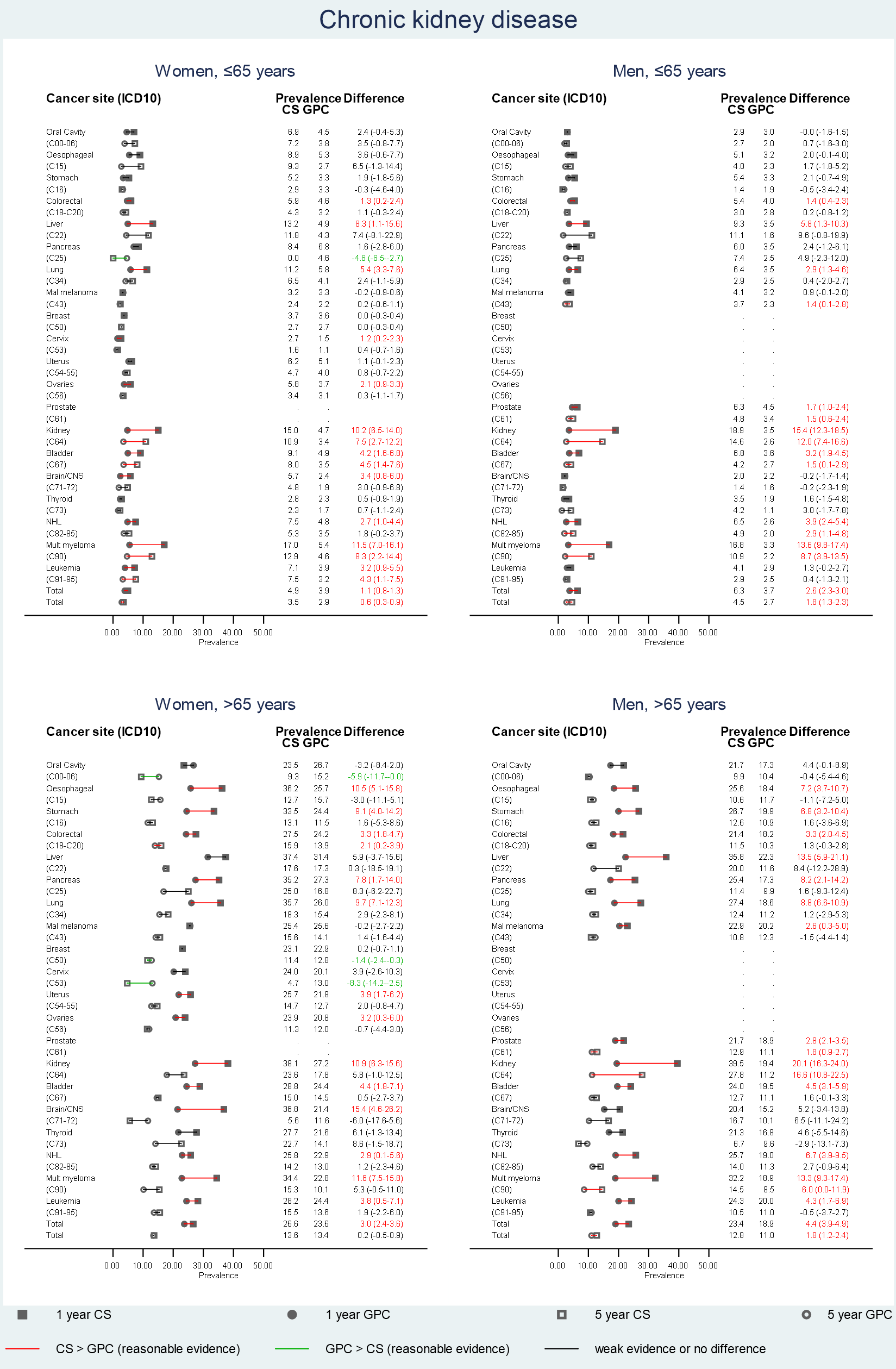

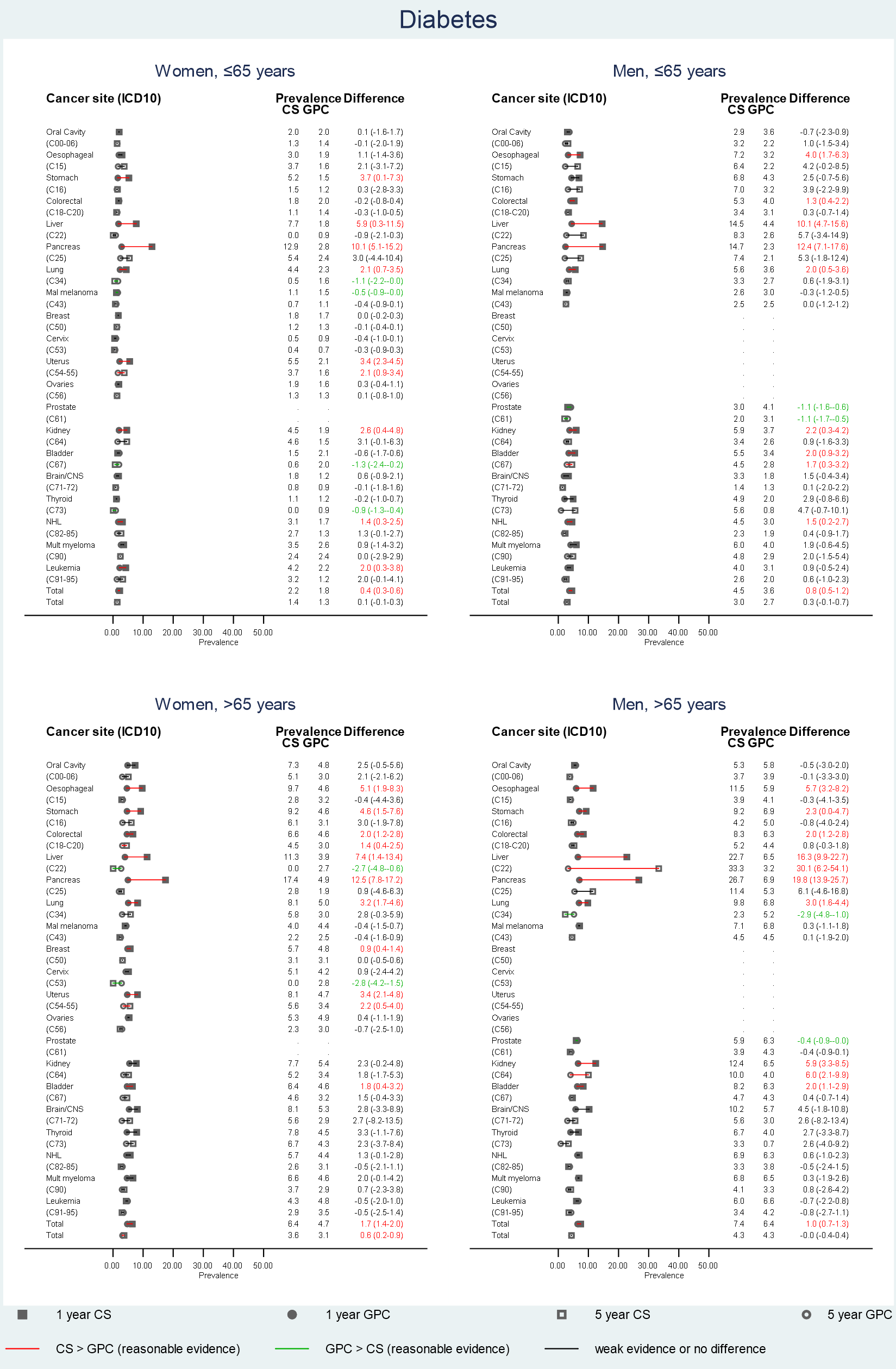

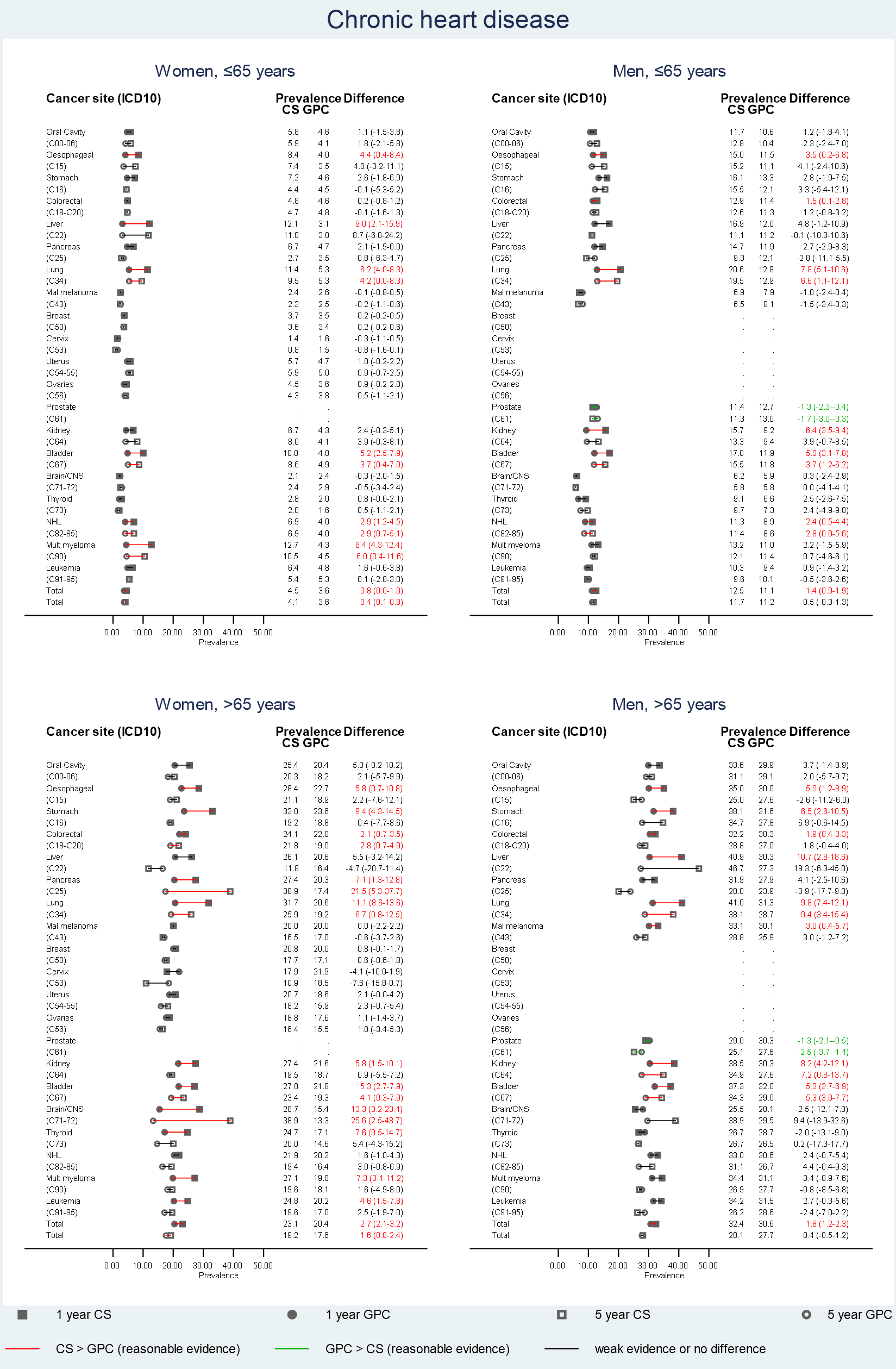

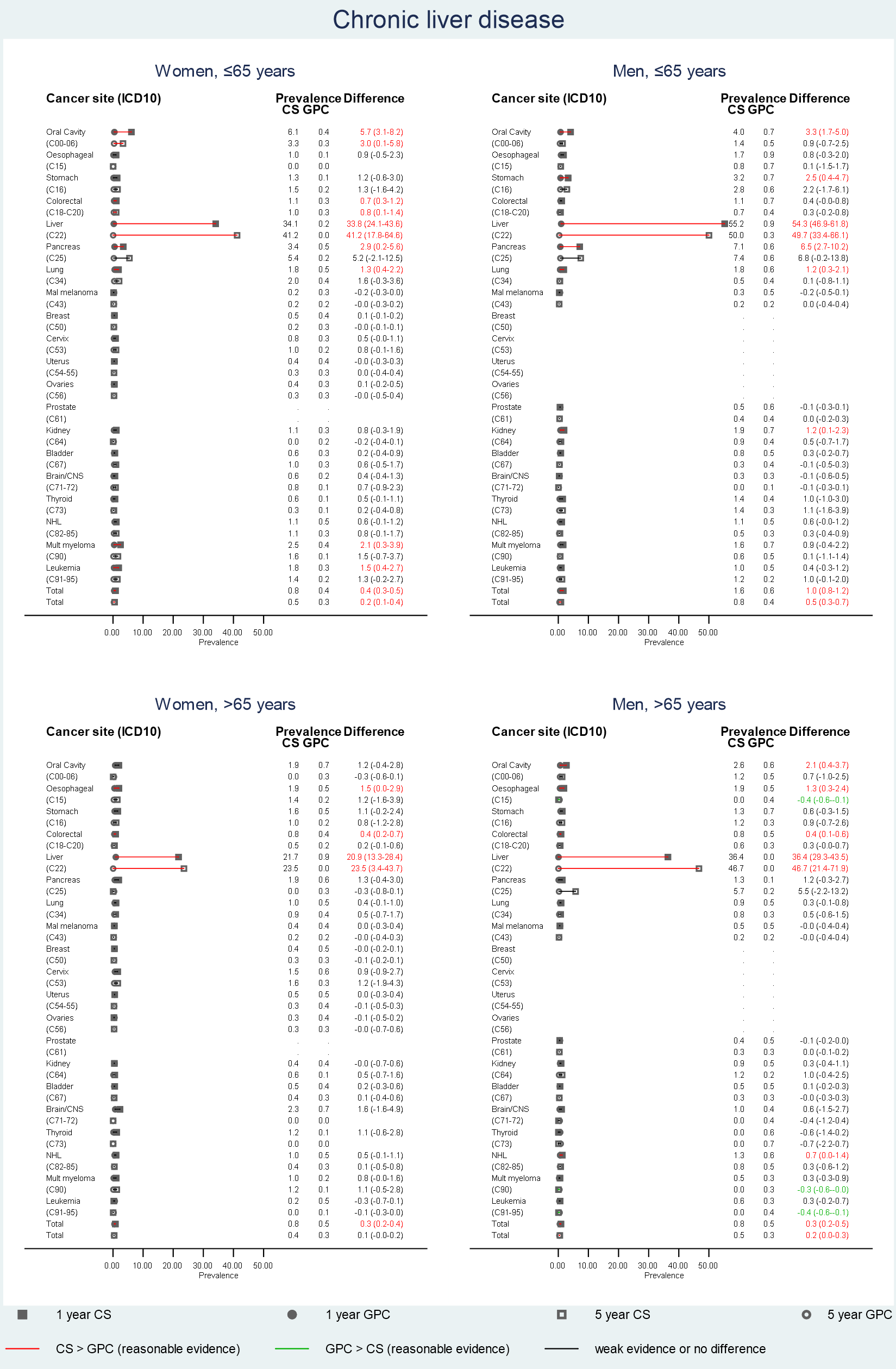

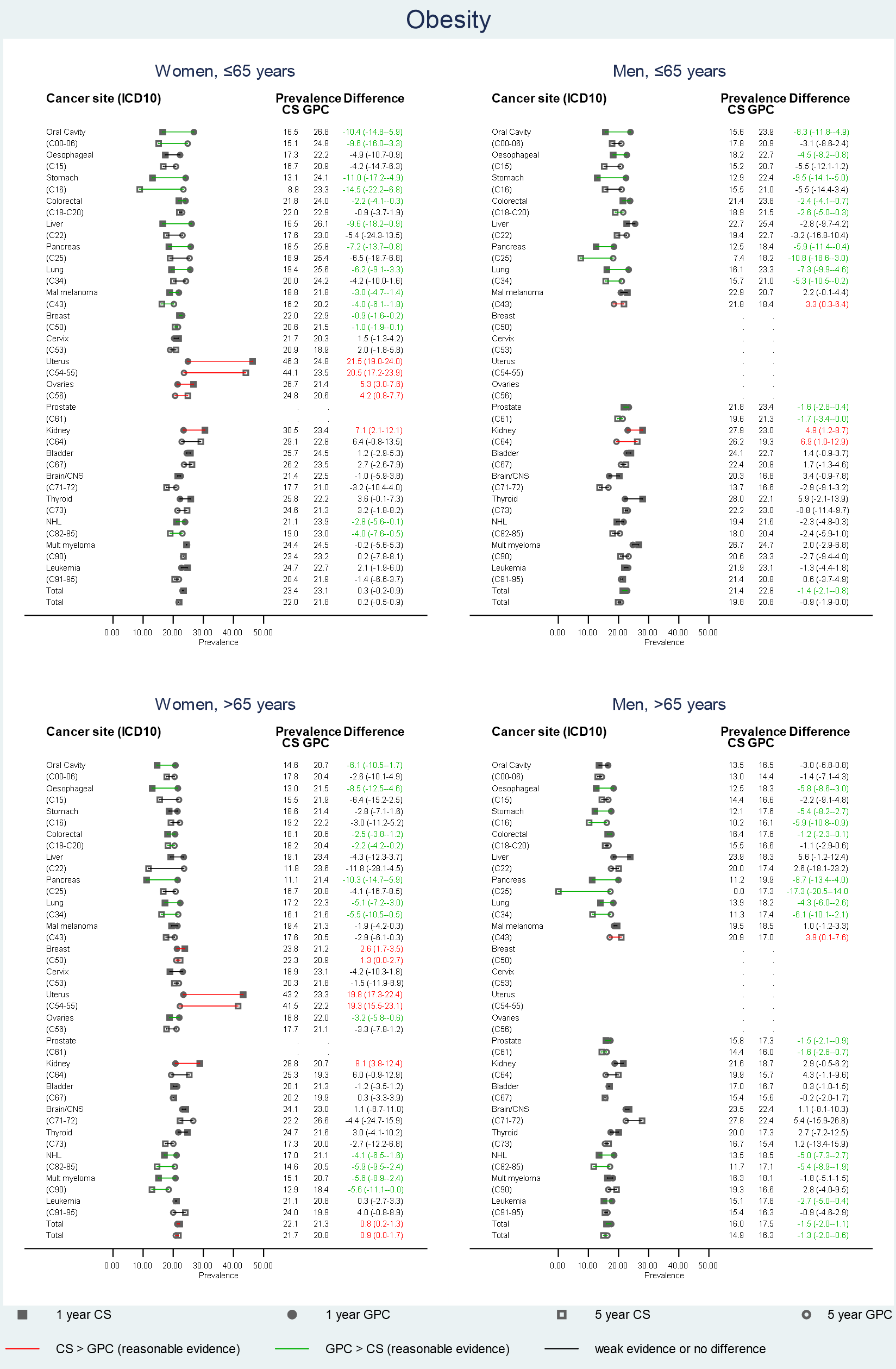
